# Receipt of COVID-19 and seasonal influenza vaccines in California (USA) during the 2021-2022 influenza season

**DOI:** 10.1101/2022.10.21.22281343

**Authors:** Kristin L. Andrejko, Jennifer F. Myers, John Openshaw, Nozomi Fukui, Sophia Li, James P. Watt, Erin L. Murray, Cora Hoover, Joseph A. Lewnard, Seema Jain, Jake M. Pry, the California COVID-19 Case-Control Study Team

## Abstract

**Background:** Despite lower circulation of influenza virus throughout 2020–2022 during the COVID-19 pandemic, seasonal influenza vaccination has remained a primary tool to reduce influenza-associated illness and death. The relationship between the decision to receive a COVID-19 vaccine and/or an influenza vaccine is not well understood.

**Methods:** We assessed predictors of receipt of 2021–2022 influenza vaccine in a secondary analysis of data from a case-control study enrolling individuals who received SARS-CoV-2 testing. We used mixed effects logistic regression to estimate factors associated with receipt of seasonal influenza vaccine. We also constructed multinomial adjusted marginal probability models of being vaccinated for COVID-19 only, seasonal influenza only, or both as compared with receipt of neither vaccination.

**Results:** Among 1261 eligible participants recruited between 22 October 2021 – 22 June 2022, 43% (545) were vaccinated with both seasonal influenza vaccine and ≥1 dose of a COVID-19 vaccine, 34% (426) received ≥1 dose of a COVID-19 vaccine only, 4% (49) received seasonal influenza vaccine only, and 19% (241) received neither vaccine. Receipt of ≥1 COVID-19 vaccine dose was associated with seasonal influenza vaccination (adjusted odds ratio [aOR]: 3.72; 95% confidence interval [CI]: 2.15–6.43); this association was stronger among participants receiving ≥1 COVID-19 booster dose (aOR=16.50 [10.10– 26.97]). Compared with participants testing negative for SARS-CoV-2 infection, participants testing positive had lower odds of receipt of 2021-2022 seasonal influenza vaccine (aOR=0.64 [0.50–0.82]).

**Conclusions:** Recipients of a COVID-19 vaccine were more likely to receive seasonal influenza vaccine during the 2021–2022 season. Factors associated with individuals’ likelihood of receiving COVID-19 and seasonal influenza vaccines will be important to account for in future studies of vaccine effectiveness against both conditions. Participants who tested positive for SARS-CoV-2 in our sample were less likely to have received seasonal influenza vaccine, suggesting an opportunity to offer influenza vaccination before or after a COVID-19 diagnosis.

## Background

Annual vaccination programs remain a critical public health tool to mitigate disease burden for seasonal influenza. Similar programs may also become an important strategy for distribution of COVID-19 vaccine doses due to waning immunity or emerging SARS-CoV-2 variants of concern in coming seasons (1, 2). Despite campaigns to improve vaccine awareness, access, and acceptance, uptake of both seasonal influenza vaccines and COVID-19 vaccines remains sub-optimal in the United States (3, 4). Vaccine hesitancy has been identified as one of the top ten threats to global health by the World Health Organization and has become increasingly prominent during the COVID-19 pandemic amid proliferation of vaccine misinformation (5). Understanding the characteristics of populations who remain unvaccinated is important to help improve coverage and ultimately reduce the burden of vaccine-preventable illness.

COVID-19 vaccine acceptance and uptake of the primary series and additional booster doses has varied throughout the pandemic (6-9). In the United States, seasonal influenza vaccine uptake increased modestly during the 2020–2021 season as compared with the 2019-2020 season (50.2% versus 48.%) (10). The relationship between individuals’ decision to receive vaccines against COVID-19 and seasonal influenza in the Unites States is unclear. Characterizing factors associated with receipt of COVID-19 or seasonal influenza vaccines may provide insights that will allow for tailoring of efforts to reach people who remain incompletely vaccinated. In addition, correlation between receipt of COVID-19 and seasonal influenza vaccines may become an important factor to adjust for in observational studies addressing the effectiveness of each vaccine against the intended pathogens (11).

We analyzed data from an ongoing state-wide case-control study which enrolled individuals receiving SARS-CoV-2 testing within the state of California. Study questionnaires collected participants’ self-reported receipt of COVID-19 and seasonal influenza vaccines between October 2021 and June 2022. These results provide insight into populations opting out of influenza or COVID-19 vaccination and can help tailor public health strategies to strengthen vaccination programs for both pathogens.

## Methods

### Study design

Data for these analyses were collected as part of an ongoing test-negative design case-control study examining risk factors for testing positive for SARS-CoV-2 virus within a general-population sample. Briefly, we enrolled California residents who received molecular tests for SARS-CoV-2 and had a positive test result (case participants) or negative test result (control participants); who had a phone number recorded in the comprehensive, statewide, reportable disease information exchange; and who consented to participate (12, 13). Participants were enrolled equally across nine multi-county regions within California (**Table S1**). Trained interviewers administered a structured questionnaire over the phone which included participants’ self-reported receipt of COVID-19 and seasonal influenza vaccines (2021–2022 season). Participants were encouraged to reference their COVID-19 vaccination card or another recall aid (calendar, e-mail reminder, text message, etc.) when providing their immunization history. Participants who reported not receiving a COVID-19 vaccine were asked to indicate why they had not yet received the vaccine as an open-ended question. Participants were additionally asked an open-ended question to indicate why they sought SARS-CoV-2 testing on the occasion which led to their study recruitment.

The study population for this analysis included participants aged ≥12 years who were enrolled between 22 October 2021 – 22 June 2022; participants aged 5-11 years were included if they enrolled on or after 29 October 2021, when this age group became eligible for COVID-19 vaccination in the United States (14). During the 2021–2022 season, 86% of all influenza vaccine doses administered were received by 22 October; thus, restricting analyses to individuals enrolled after this point in time was expected to mitigate the risk of misclassifying participants’ seasonal influenza vaccination status (15).

### Exposures

The primary exposures of interest were receipt of COVID-19 vaccine(s) and 2021–2022 seasonal influenza vaccine. We defined COVID-19 vaccination status as a categorical variable denoting whether the participant had initiated a primary series of COVID-19 vaccine doses (excluding booster doses); one or more booster doses; or no receipt of any COVID-19 vaccine doses. Participants were categorized as having initiated a primary series if they were vaccinated with one dose of Ad.26.COV2.S [Jansen] or 1–2 doses of BNT162b2 [Pfizer/BioNTech] or mRNA-1273 [Moderna]. Participants were categorized as “boosted” if they received a second dose of any COVID-19 vaccine product after one dose of Ad.26.COV2.S [Jansen], or a third or fourth dose after receipt of two doses of BNT162b2 [Pfizer/BioNTech] or mRNA-1273 [Moderna]. We considered individuals to have received 2021–2022 seasonal influenza vaccine if they reported receipt of any influenza vaccine dose during 1 August 2021 – 22 June 2022.

### Statistical Analysis

We first estimated the association between receipt of COVID-19 and seasonal influenza vaccination using mixed effects logistic regression among all study participants. Models allowed for random effects for week of test and region. Potential predictors defined as model covariates included participants’ age, sex assigned at birth, region of residence, interview date, race/ethnicity, SARS-CoV-2 infection status at time of interview, and self-reported history of co-morbid conditions. To further understand how receipt of each vaccine related to other risk-reducing behaviors, we included a variable indicating self-reported use of face masks in public indoor settings during the two weeks before individuals were tested, defined as any mask use versus no mask use. We also included a variable indicating any attendance at social gatherings in the two weeks preceding SARS-CoV-2 testing. We further conducted subgroup analyses evaluating these factors as predictors of seasonal influenza vaccination among participants who reported receipt of any COVID-19 vaccine doses, and who did not report receipt of any COVID-19 vaccine doses.

Last, to define how these variables were associated with each potential vaccination status, we used adjusted multinomial regression to estimate the marginal probabilities of being unvaccinated, vaccinated for both COVID-19 and influenza, or vaccinated for only one of COVID-19 or seasonal influenza. Here, we defined COVID-19 vaccination as receipt of ≥1 dose of a COVID-19 vaccine. Probabilities estimated by this approach were not expected to allow us to infer population-wide vaccine uptake because our sample was recruited based on SARS-CoV-2 test outcome; nonetheless, this analysis was expected to identify factors distinguishing recipients of one or both vaccines from recipients of neither vaccine.

All analysis were completed using Stata 17 (StataCorp LC, College Station, TX, USA).

### Ethics

The study protocol was approved by the State of California, Health and Human Services Agency, Committee for the Protection of Human Subjects (Project Number: 2021-034).

## Results

### Descriptive characteristics of the study population

A total of 1261 participants were included in the analysis with 56% identifying as female; the median age was 35 years (interquartile range [IQR]: 24-53 years). Among all 1261 participants, 43% (545) received seasonal influenza vaccine during the 2021–22 season and ≥1 dose of a COVID-19 vaccine at any time; 34% (426) received ≥1 dose of a COVID-19 vaccine but no seasonal influenza vaccine, 4% (49) received seasonal influenza vaccine only, and 19% (241) received neither vaccine (**Table 1; Table S2**). The majority of participants who received seasonal influenza vaccine only were aged 5-17 (71%; 35/49), consistent with the timeline of COVID-19 vaccine becoming available to children aged 5–11 years only from 29 October 2021 onward. Among 971 participants who received ≥1 dose COVID-19 vaccine by the time of their interview, 53% (511) received ≥1 booster dose (**Table S3)**. Only 27 participants who had received ≥1 dose of an mRNA-based COVID-19 vaccine had not yet received a second dose. Among 511 participants who received any booster dose, the majority received only one booster dose (490, 95.9%) and 21 (4.1%) received two booster doses (**Table S4**). Participants who received at least one booster dose were older than 11 years of age and most did not report a history of co-morbid conditions (66.9%, 327/490); participants who received two booster doses were mostly adults over the age of 50 (90%; 19/21) and 60% (12/21) reported having at least one co-morbidity (**Table S4**).

**Table 1:**
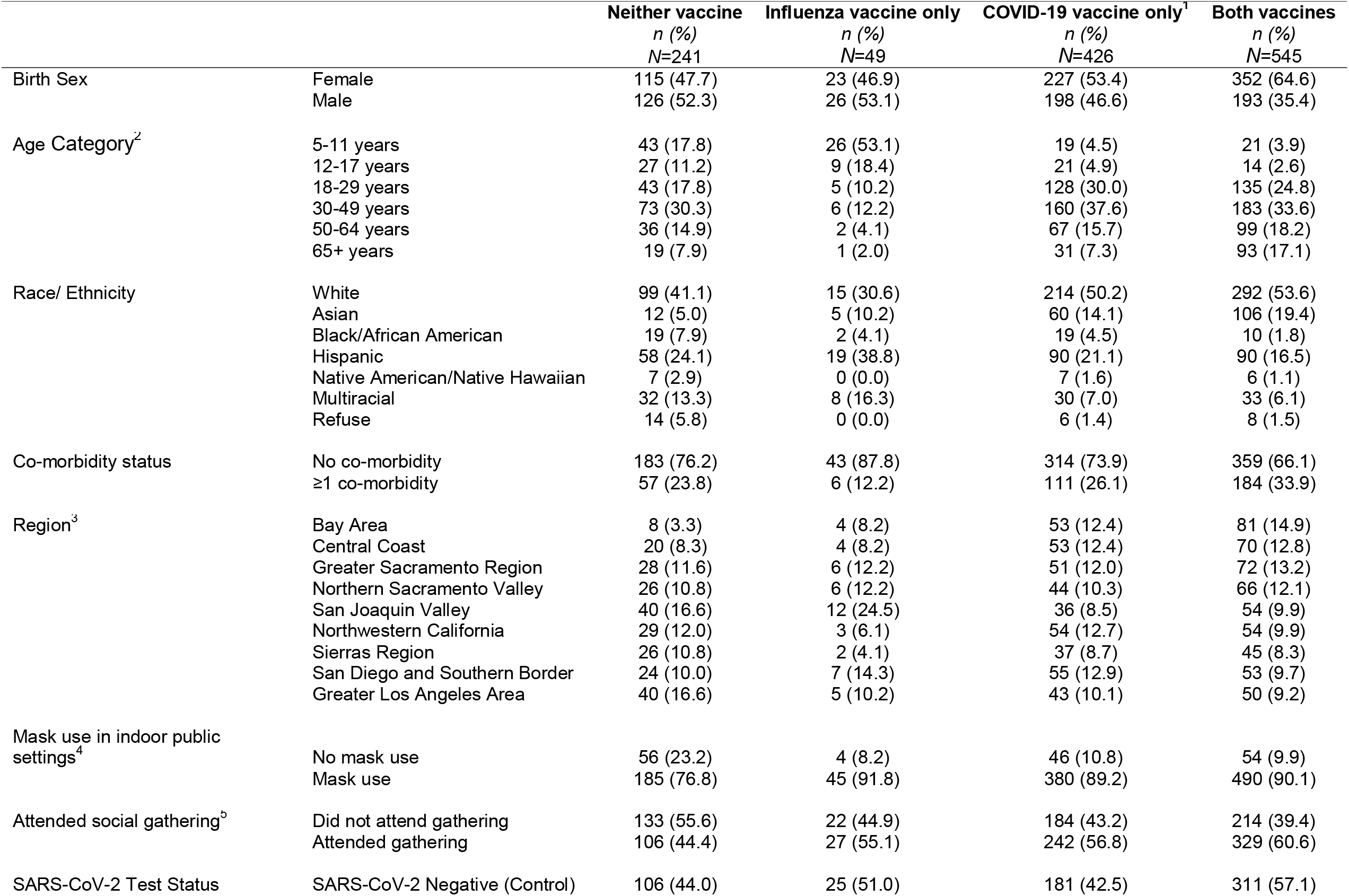
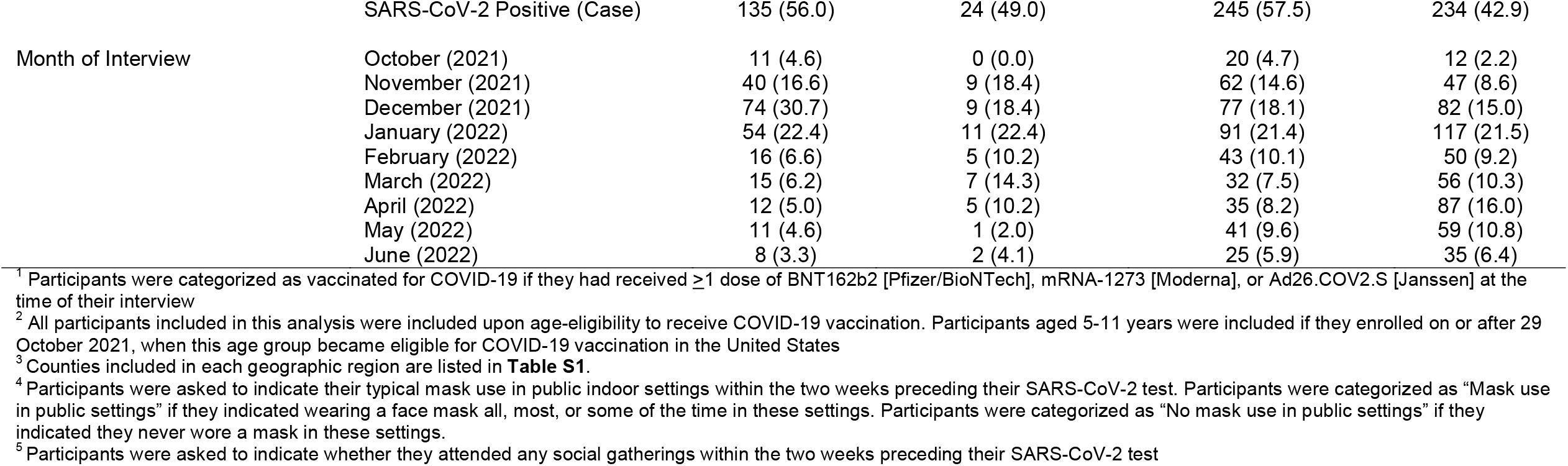
Population characteristics by seasonal influenza vaccination (2021-2022) and receipt of one or more doses of a COVID-19 vaccine (*N=*1261).

|  |  | Neither vaccine<br>*n* (%)<br>N=241 | Influenza vaccine only<br>*n* (%)<br>N=49 | COVID-19 vaccine only<sup>1</sup><br>*n* (%)<br>N=426 | Both vaccines<br>*n* (%)<br>N=545 |
| --- | --- | --- | --- | --- | --- |
| Birth Sex | Female | 115 (47.7) | 23 (46.9) | 227 (53.4) | 352 (64.6) |
|  | Male | 126 (52.3) | 26 (53.1) | 198 (46.6) | 193 (35.4) |
| Age Category<sup>2</sup> | 5-11 years | 43 (17.8) | 26 (53.1) | 19 (4.5) | 21 (3.9) |
|  | 12-17 years | 27 (11.2) | 9 (18.4) | 21 (4.9) | 14 (2.6) |
|  | 18-29 years | 43 (17.8) | 5 (10.2) | 128 (30.0) | 135 (24.8) |
|  | 30-49 years | 73 (30.3) | 6 (12.2) | 160 (37.6) | 183 (33.6) |
|  | 50-64 years | 36 (14.9) | 2 (4.1) | 67 (15.7) | 99 (18.2) |
|  | 65+ years | 19 (7.9) | 1 (2.0) | 31 (7.3) | 93 (17.1) |
| Race/ Ethnicity | White | 99 (41.1) | 15 (30.6) | 214 (50.2) | 292 (53.6) |
|  | Asian | 12 (5.0) | 5 (10.2) | 60 (14.1) | 106 (19.4) |
|  | Black/African American | 19 (7.9) | 2 (4.1) | 19 (4.5) | 10 (1.8) |
|  | Hispanic | 58 (24.1) | 19 (38.8) | 90 (21.1) | 90 (16.5) |
|  | Native American/Native Hawaiian | 7 (2.9) | 0 (0.0) | 7 (1.6) | 6 (1.1) |
|  | Multiracial | 32 (13.3) | 8 (16.3) | 30 (7.0) | 33 (6.1) |
|  | Refuse | 14 (5.8) | 0 (0.0) | 6 (1.4) | 8 (1.5) |
| Co-morbidity status | No co-morbidity | 183 (76.2) | 43 (87.8) | 314 (73.9) | 359 (66.1) |
|  | ≥1 co-morbidity | 57 (23.8) | 6 (12.2) | 111 (26.1) | 184 (33.9) |
| Region<sup>3</sup> | Bay Area | 8 (3.3) | 4 (8.2) | 53 (12.4) | 81 (14.9) |
|  | Central Coast | 20 (8.3) | 4 (8.2) | 53 (12.4) | 70 (12.8) |
|  | Greater Sacramento Region | 28 (11.6) | 6 (12.2) | 51 (12.0) | 72 (13.2) |
|  | Northern Sacramento Valley | 26 (10.8) | 6 (12.2) | 44 (10.3) | 66 (12.1) |
|  | San Joaquin Valley | 40 (16.6) | 12 (24.5) | 36 (8.5) | 54 (9.9) |
|  | Northwestern California | 29 (12.0) | 3 (6.1) | 54 (12.7) | 54 (9.9) |
|  | Sierras Region | 26 (10.8) | 2 (4.1) | 37 (8.7) | 45 (8.3) |
|  | San Diego and Southern Border | 24 (10.0) | 7 (14.3) | 55 (12.9) | 53 (9.7) |
|  | Greater Los Angeles Area | 40 (16.6) | 5 (10.2) | 43 (10.1) | 50 (9.2) |
| Mask use in indoor public settings<sup>4</sup> | No mask use | 56 (23.2) | 4 (8.2) | 46 (10.8) | 54 (9.9) |
|  | Mask use | 185 (76.8) | 45 (91.8) | 380 (89.2) | 490 (90.1) |
| Attended social gathering<sup>5</sup> | Did not attend gathering | 133 (55.6) | 22 (44.9) | 184 (43.2) | 214 (39.4) |
|  | Attended gathering | 106 (44.4) | 27 (55.1) | 242 (56.8) | 329 (60.6) |
| SARS-CoV-2 Test Status | SARS-CoV-2 Negative (Control) | 106 (44.0) | 25 (51.0) | 181 (42.5) | 311 (57.1) |

|  | SARS-CoV-2 Positive (Case) | 135 (56.0) | 24 (49.0) | 245 (57.5) | 234 (42.9) |
| --- | --- | --- | --- | --- | --- |
| Month of Interview | October (2021) | 11 (4.6) | 0 (0.0) | 20 (4.7) | 12 (2.2) |
|  | November (2021) | 40 (16.6) | 9 (18.4) | 62 (14.6) | 47 (8.6) |
|  | December (2021) | 74 (30.7) | 9 (18.4) | 77 (18.1) | 82 (15.0) |
|  | January (2022) | 54 (22.4) | 11 (22.4) | 91 (21.4) | 117 (21.5) |
|  | February (2022) | 16 (6.6) | 5 (10.2) | 43 (10.1) | 50 (9.2) |
|  | March (2022) | 15 (6.2) | 7 (14.3) | 32 (7.5) | 56 (10.3) |
|  | April (2022) | 12 (5.0) | 5 (10.2) | 35 (8.2) | 87 (16.0) |
|  | May (2022) | 11 (4.6) | 1 (2.0) | 41 (9.6) | 59 (10.8) |
|  | June (2022) | 8 (3.3) | 2 (4.1) | 25 (5.9) | 35 (6.4) |

<sup>1</sup> Participants were categorized as vaccinated for COVID-19 if they had received $\geq 1$ dose of BNT162b2 [Pfizer/BioNTech], mRNA-1273 [Moderna], or Ad26.COV2.S [Janssen] at the time of their interview
<sup>2</sup> All participants included in this analysis were included upon age-eligibility to receive COVID-19 vaccination. Participants aged 5-11 years were included if they enrolled on or after 29 October 2021, when this age group became eligible for COVID-19 vaccination in the United States
<sup>3</sup> Counties included in each geographic region are listed in **Table S1**.
<sup>4</sup> Participants were asked to indicate their typical mask use in public indoor settings within the two weeks preceding their SARS-CoV-2 test. Participants were categorized as "Mask use in public settings" if they indicated wearing a face mask all, most, or some of the time in these settings. Participants were categorized as "No mask use in public settings" if they indicated they never wore a mask in these settings.
<sup>5</sup> Participants were asked to indicate whether they attended any social gatherings within the two weeks preceding their SARS-CoV-2 test

Presence of COVID-19 symptoms was the most cited reason for testing for all participants, regardless of history of COVID-19 and seasonal influenza vaccine receipt, followed by contact with an individual known or suspected to have been infected with SARS-CoV-2, either in the household or in other settings (**Table 2**). When further stratified by SARS-CoV-2 test status, this observation held among the participants who tested positive for SARS-CoV-2. However, among participants who tested negative for SARS-CoV-2, the most common reason for testing was screening for work or school, regardless of vaccination status (**Table S5)**.

**Table 2.** Reason for SARS-CoV-2 testing stratified by receipt of COVID-19 and influenzas vaccination

| *Reason for SARS-CoV-2 testing<sup>2</sup>* | **Neither vaccine**<br>*n (%)*<br>*N = 241* | **Influenza vaccine only**<br>*n (%)*<br>*N = 49* | **COVID-19 vaccine only<sup>1</sup>**<br>*n (%)*<br>*N = 426* | **Both vaccines**<br>*n (%)*<br>*N = 545* |
| --- | --- | --- | --- | --- |
| Experiencing COVID-19 symptoms | 99 (41.1) | 21 (42.9) | 198 (46.5) | 188 (34.5) |
| Household SARS-CoV-2 exposure | 12 (5.0) | 2 (4.1) | 12 (2.8) | 15 (2.8) |
| Other SARS-CoV-2 exposure | 57 (23.7) | 13 (26.5) | 113 (26.5) | 134 (24.6) |
| Required screening test for work/school | 57 (23.7) | 8 (16.3) | 66 (15.5) | 128 (23.5) |
| Curious to see if infected | 37 (15.4) | 7 (14.3) | 67 (15.7) | 106 (19.4) |
| Medical procedure or admission to hospital | 21 (8.7) | 7 (14.3) | 31 (7.3) | 66 (12.1) |
| Travel requirement | 9 (3.7) | 1 (2.0) | 23 (5.4) | 27 (5.0) |
| Public health recommendation to get tested | 2 (0.8) | 0 (0.0) | 4 (0.9) | 4 (0.7) |

<sup>1</sup>Since interviewers indicated all reasons listed by participants, and some participants refused to respond to the question, reasons will not sum to the total sample size.
<sup>2</sup>Participants were categorized as vaccinated for COVID-19 if they had received $\geq 1$ dose of BNT162b2 [Pfizer/BioNTech], mRNA-1273 [Moderna], or Ad26.COV2.S [Janssen] at the time of their interview

### Association of seasonal influenza vaccination with COVID-19 vaccination

Initiation of the COVID-19 vaccine series was associated with 3.72-fold (95%CI: 2.15–6.43) higher adjusted odds of receipt of seasonal influenza vaccine, compared with not receiving any COVID-19 vaccine doses (**Table 3**). Participants who received a COVID-19 vaccine booster dose had 16.50-fold (95%CI: 10.10–26.97) higher odds of receiving seasonal influenza vaccine than participants who received no COVID-19 vaccine doses.

**Table 3:**
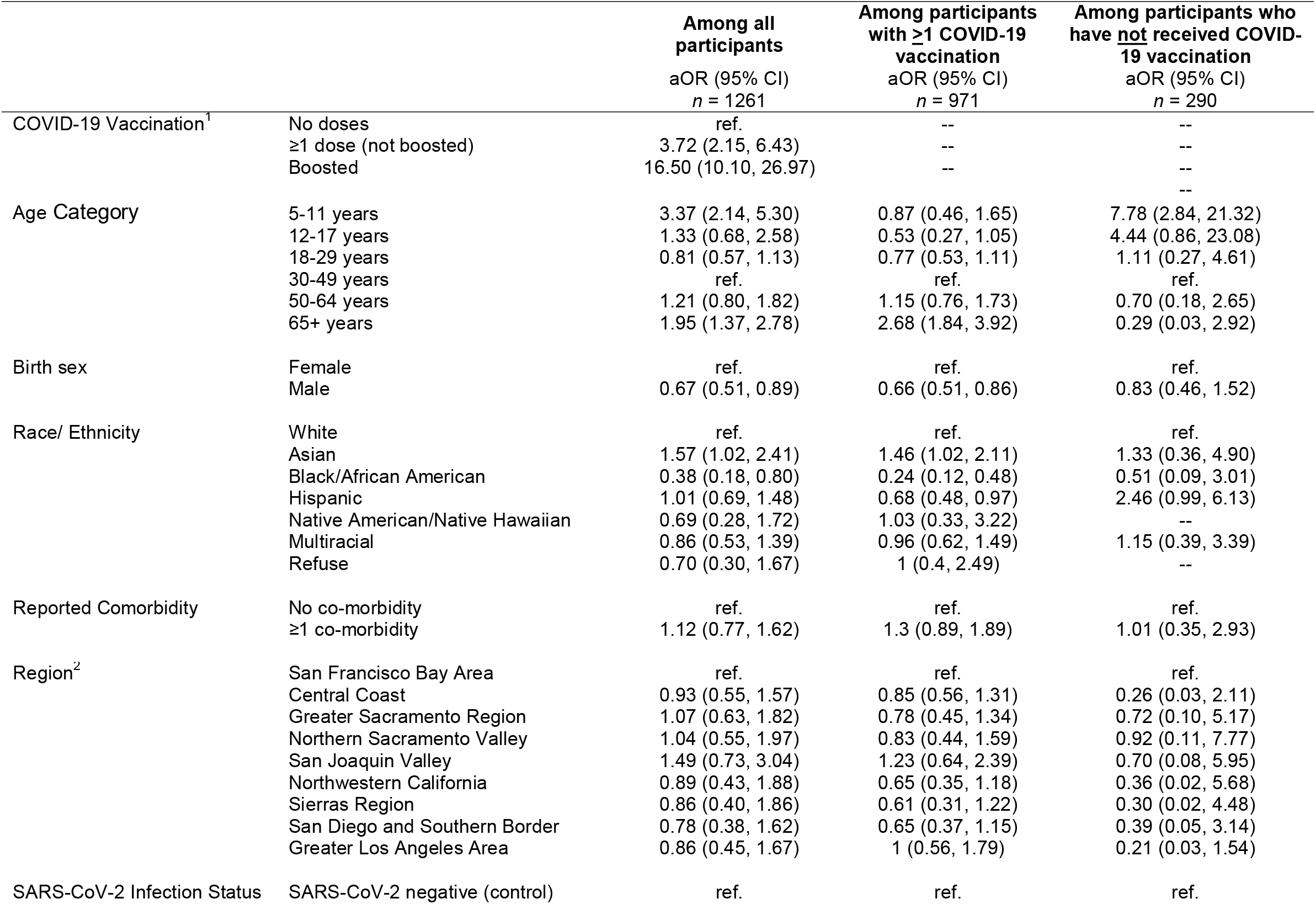
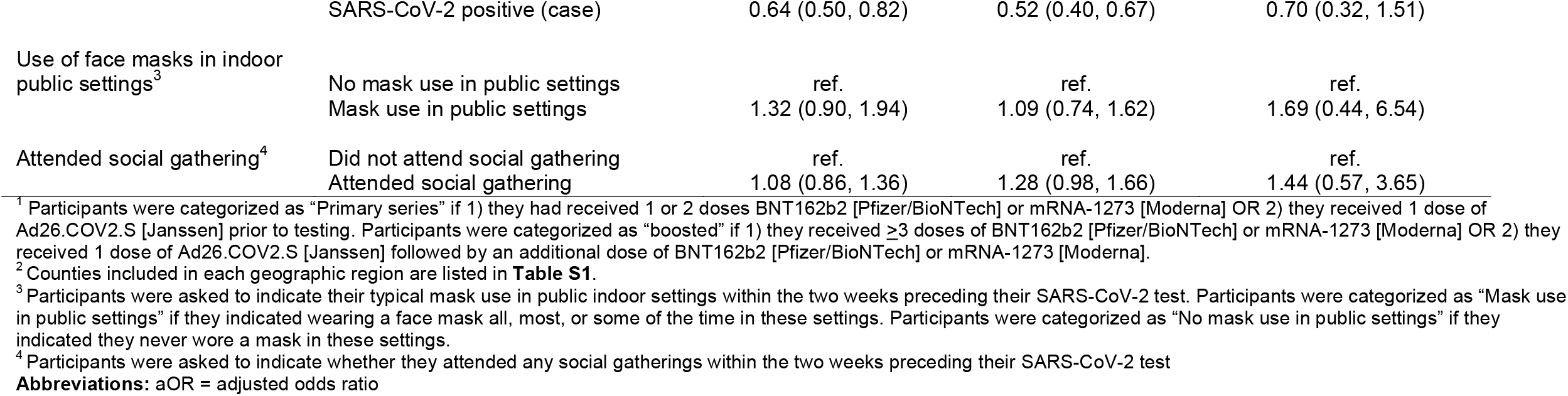
Adjusted Odds Ratios for Influenza Vaccination Status among all participants and among participants with at least one COVID-19 vaccine dose (N=1261).

|  |  | Among all<br>participants<br>aOR (95% CI)<br>n = 1261 | Among participants<br>with ≥1 COVID-19<br>vaccination<br>aOR (95% CI)<br>n = 971 | Among participants who<br>have <u>not</u> received COVID-19 vaccination<br>aOR (95% CI)<br>n = 290 |
| --- | --- | --- | --- | --- |
| COVID-19 Vaccination<sup>1</sup> | No doses | ref. | -- | -- |
|  | ≥1 dose (not boosted) | 3.72 (2.15, 6.43) | -- | -- |
|  | Boosted | 16.50 (10.10, 26.97) | -- | -- |
|  |  |  |  | -- |
| Age Category | 5-11 years | 3.37 (2.14, 5.30) | 0.87 (0.46, 1.65) | 7.78 (2.84, 21.32) |
|  | 12-17 years | 1.33 (0.68, 2.58) | 0.53 (0.27, 1.05) | 4.44 (0.86, 23.08) |
|  | 18-29 years | 0.81 (0.57, 1.13) | 0.77 (0.53, 1.11) | 1.11 (0.27, 4.61) |
|  | 30-49 years | ref. | ref. | ref. |
|  | 50-64 years | 1.21 (0.80, 1.82) | 1.15 (0.76, 1.73) | 0.70 (0.18, 2.65) |
|  | 65+ years | 1.95 (1.37, 2.78) | 2.68 (1.84, 3.92) | 0.29 (0.03, 2.92) |
| Birth sex | Female | ref. | ref. | ref. |
|  | Male | 0.67 (0.51, 0.89) | 0.66 (0.51, 0.86) | 0.83 (0.46, 1.52) |
| Race/ Ethnicity | White | ref. | ref. | ref. |
|  | Asian | 1.57 (1.02, 2.41) | 1.46 (1.02, 2.11) | 1.33 (0.36, 4.90) |
|  | Black/African American | 0.38 (0.18, 0.80) | 0.24 (0.12, 0.48) | 0.51 (0.09, 3.01) |
|  | Hispanic | 1.01 (0.69, 1.48) | 0.68 (0.48, 0.97) | 2.46 (0.99, 6.13) |
|  | Native American/Native Hawaiian | 0.69 (0.28, 1.72) | 1.03 (0.33, 3.22) | -- |
|  | Multiracial | 0.86 (0.53, 1.39) | 0.96 (0.62, 1.49) | 1.15 (0.39, 3.39) |
|  | Refuse | 0.70 (0.30, 1.67) | 1 (0.4, 2.49) | -- |
| Reported Comorbidity | No co-morbidity | ref. | ref. | ref. |
|  | ≥1 co-morbidity | 1.12 (0.77, 1.62) | 1.3 (0.89, 1.89) | 1.01 (0.35, 2.93) |
| Region<sup>2</sup> | San Francisco Bay Area | ref. | ref. | ref. |
|  | Central Coast | 0.93 (0.55, 1.57) | 0.85 (0.56, 1.31) | 0.26 (0.03, 2.11) |
|  | Greater Sacramento Region | 1.07 (0.63, 1.82) | 0.78 (0.45, 1.34) | 0.72 (0.10, 5.17) |
|  | Northern Sacramento Valley | 1.04 (0.55, 1.97) | 0.83 (0.44, 1.59) | 0.92 (0.11, 7.77) |
|  | San Joaquin Valley | 1.49 (0.73, 3.04) | 1.23 (0.64, 2.39) | 0.70 (0.08, 5.95) |
|  | Northwestern California | 0.89 (0.43, 1.88) | 0.65 (0.35, 1.18) | 0.36 (0.02, 5.68) |
|  | Sierras Region | 0.86 (0.40, 1.86) | 0.61 (0.31, 1.22) | 0.30 (0.02, 4.48) |
|  | San Diego and Southern Border | 0.78 (0.38, 1.62) | 0.65 (0.37, 1.15) | 0.39 (0.05, 3.14) |
|  | Greater Los Angeles Area | 0.86 (0.45, 1.67) | 1 (0.56, 1.79) | 0.21 (0.03, 1.54) |
| SARS-CoV-2 Infection Status | SARS-CoV-2 negative (control) | ref. | ref. | ref. |
|  | SARS-CoV-2 positive (case) | 0.64 (0.50, 0.82) | 0.52 (0.40, 0.67) | 0.70 (0.32, 1.51) |
| Use of face masks in indoor public settings<sup>3</sup> | No mask use in public settings | ref. | ref. | ref. |
|  | Mask use in public settings | 1.32 (0.90, 1.94) | 1.09 (0.74, 1.62) | 1.69 (0.44, 6.54) |
| Attended social gathering<sup>4</sup> | Did not attend social gathering | ref. | ref. | ref. |
|  | Attended social gathering | 1.08 (0.86, 1.36) | 1.28 (0.98, 1.66) | 1.44 (0.57, 3.65) |

<sup>1</sup> Participants were categorized as “Primary series” if 1) they had received 1 or 2 doses BNT162b2 [Pfizer/BioNTech] or mRNA-1273 [Moderna] OR 2) they received 1 dose of Ad26.COV2.S [Janssen] prior to testing. Participants were categorized as “boosted” if 1) they received $\geq 3$ doses of BNT162b2 [Pfizer/BioNTech] or mRNA-1273 [Moderna] OR 2) they received 1 dose of Ad26.COV2.S [Janssen] followed by an additional dose of BNT162b2 [Pfizer/BioNTech] or mRNA-1273 [Moderna].
<sup>2</sup> Counties included in each geographic region are listed in **Table S1**.
<sup>3</sup> Participants were asked to indicate their typical mask use in public indoor settings within the two weeks preceding their SARS-CoV-2 test. Participants were categorized as “Mask use in public settings” if they indicated wearing a face mask all, most, or some of the time in these settings. Participants were categorized as “No mask use in public settings” if they indicated they never wore a mask in these settings.
<sup>4</sup> Participants were asked to indicate whether they attended any social gatherings within the two weeks preceding their SARS-CoV-2 test
**Abbreviations:** aOR = adjusted odds ratio

As compared with participants aged 30-49 years, participants aged 5-11 years had 3.37 (95% CI: 2.15– 6.43) fold higher adjusted odds of receiving seasonal influenza vaccine. Participants aged >65 years had 1.95 (95%CI: 1.37–2.78) fold higher adjusted odds of receiving seasonal influenza vaccine than participants 30-49 years old. Males had lower adjusted odds of receiving influenza vaccination than females (aOR = 0.67 [95%CI: 0.51–0.89]). Asian participants had 1.57 (95%CI: 1.08–2.41) fold higher odds of receiving seasonal influenza vaccine than White participants, while Black/African American participants had lower odds of receipt of seasonal influenza vaccine than White participants (aOR=0.38 (0.18-0.80).

Estimates did not suggest differences in odds of receiving seasonal influenza vaccine among participants who did or did not attend social gatherings. Adjusted odds of receiving seasonal influenza vaccine were 1.32 (0.90-1.94) fold higher among participants who reported any use of face masks in indoor public settings in the prior two weeks, as compared with participants who reported not using face masks. We did not observe differences in seasonal influenza vaccine uptake according to participants’ self-reported co-morbidities or region of residence. Participants who had recently (previous ≤7 days) tested positive for SARS-CoV-2 had 0.64-fold (95%CI: 0.50–0.82) lower odds of having received seasonal influenza vaccine as compared with participants who tested negative.

We also assessed predictors of seasonal influenza vaccine receipt among the 971 participants who received at least one dose of a COVID-19 vaccine. Participants aged ≥65 years had 2.68-fold (95% CI: 1.84–3.92) higher adjusted odds of receiving seasonal influenza vaccine as compared with participants aged 30-49 years (**Table 3; Figure S1**). Males had lower adjusted odds of seasonal influenza vaccine receipt than females (aOR = 0.66; 95%CI: 0.51–0.86). While Hispanic and Black/African American participants had lower adjusted odds of seasonal influenza vaccine receipt as compared to White participants (aOR = 0.68 [0.48–0.97] for Hispanic; aOR = 0.24 [0.12–0.48] for Black/African American), participants identifying as Asian had higher adjusted odds of seasonal influenza vaccine receipt (aOR = 1.46 [1.02–2.11]). Participants who tested positive for SARS-CoV-2 had lower adjusted odds (aOR = 0.52 [0.40–0.67]) of receipt of seasonal influenza vaccine than participants who tested negative. Seasonal influenza vaccination among participants who received at least one COVID-19 vaccine dose was not associated with self-reported comorbidities, region of residence, or use of face masks in public indoor settings.

Among the 290 participants who did not receive any COVID-19 vaccine doses, younger age (5-11 years and 12-17 years) was a strong predictor of seasonal influenza vaccine receipt (aOR = 7.78 [2.84–21.32] at ages 5-11 years vs. 30-49 years; aOR = 4.44 [0.86–23.08] at ages 12–17 vs. 30-49 years; **Table 3**). We did not find an association of birth sex with seasonal influenza vaccination in this stratum (aOR = 0.83 [0.46-1.52]). Participants of Hispanic ethnicity had the highest adjusted odds of receiving only influenza vaccination (aOR = 2.46 [0.99–6.13] for Hispanic vs. White participants). Adjusted odds of seasonal influenza vaccination were 1.44 (0.57-3.65) and 1.69 (0.44-6.54) fold higher among participants who reported attending social gatherings and wearing masks in indoor public settings in the preceding two weeks, respectively, in comparison to participants who did not attend social gatherings and who did not wear face masks.

Age was a strong predictor of receiving seasonal influenza and COVID-19 vaccines. For participants aged ≥65 years, the probability of receipt of both seasonal influenza and COVID-19 vaccines reached 62.8% (95% CI: 54.8, 70.9%), while the highest probability of receiving neither COVID-19 or influenza vaccine was observed among participants aged 5-11 years (38.3% [28.7–47.8%]; **Fig 1A**). Participants aged 18-29 had the highest probability of being vaccinated against COVID-19 only. Participants identifying as Asian had the highest probability of receiving both vaccines at 56.9% (95% CI: 50.3–63.6%) while, in contrast, participants identifying as Black/African American had the highest probability of receiving neither COVID-19 nor influenza vaccination at 41.1% (95% CI: 28.3–53.8%). Participants residing in San Francisco Bay Area counties had the highest probability of receiving both vaccines at 53.6% (95% CI: 45.9–61.4%) (**Fig 1D**). Residents of counties within the San Joaquin Valley region had the highest probability of receiving neither COVID-19 or influenza vaccination at 24.5% (95% CI: 18.0–31.1%).

**Figure 1:**
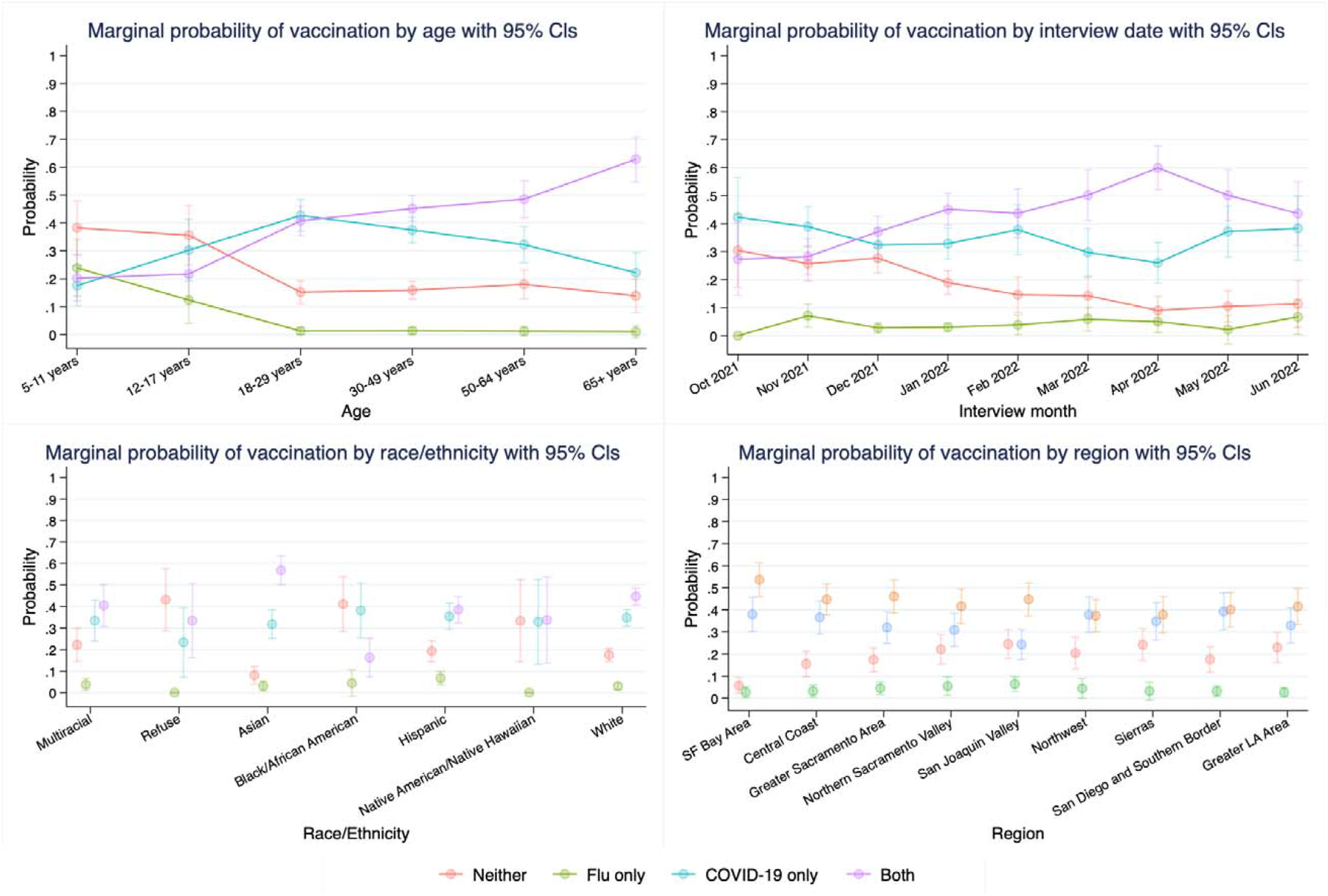
Marginal probability of receipt of an influenza vaccination. Estimates are derived from multinomial models and are presented by A) age category B) SARS-CoV-2 test month associated with parent study enrollment C) race/ethnicity D) region. Abbreviations: CI – confidence interval

**Abbreviations:** CI – confidence interval

### Attitudes toward COVID-19 vaccination

We then assessed whether the stated reasons for not receiving COVID-19 vaccination differed in association with a participants’ receipt of influenza vaccination. Among the 231 participants who received neither seasonal influenza nor COVID-19 vaccines, the primary reasons for not receiving COVID-19 vaccination were belief in the right to choose whether to be vaccinated (26%; 61), a desire to wait for more research (19%; 44), and/or fear of short-term side effects (13%; 32) (**Table 4**). Among the 49 participants who had not received COVID-19 vaccination but received seasonal influenza vaccination, the top three reasons for not receiving COVID-19 vaccination included the right to choose (22.9%; 11), need for more research (16.4%, 8), and/or side effects (10.4%, 5). None of the participants who did not receive COVID-19 vaccination but were vaccinated for seasonal influenza cited concerns about COVID-19 vaccine safety, religious reason(s), previously or currently infected with SARS-CoV-2, general vaccine safety, or distrust in the government and/or medical institutions.

**Table 4.** Reasons for not receiving COVID-19 vaccination by seasonal influenza vaccination status

| *Reason for not receiving COVID-19 vaccination<sup>1</sup>* | **No influenza**<br>**vaccination** | **Influenza**<br>**vaccinated** | *p-value<sup>2</sup>* |
| --- | --- | --- | --- |
|  | n (%)<br>N = 231 | n (%)<br>N = 49 |  |
| Right to choose whether to vaccinate | 61 (25.6) | 11 (22.9) | 0.855 |
| More research needed | 44 (18.5) | 8 (16.7) | 0.840 |
| Side effects | 32 (13.4) | 5 (10.4) | 0.813 |
| Long-term side effects | 27 (11.3) | 2 (4.2) | 0.189 |
| Not important for me | 21 (8.8) | 2 (4.2) | 0.389 |
| Concerns about pregnancy | 18 (7.6) | 1 (2.1) | 0.216 |
| Not enough information | 15 (6.3) | 3 (6.3) | 1.000 |
| Concerns about safety for children | 14 (5.9) | 0 (0) | 0.137 |
| Concerned COVID-19 vaccine safety | 13 (5.5) | 0 (0) | 0.135 |
| Not at high risk for SARS-CoV-2 infection | 12 (5) | 1 (2.1) | 0.702 |
| Vaccines are generally not safe | 10 (4.2) | 1 (2.1) | 0.697 |
| Religious reason(s) | 10 (4.2) | 0 (0) | 0.222 |
| Medical condition(s) | 9 (3.8) | 6 (12.5) | 0.025 |
| Prior/current COVID-19 infection | 8 (3.4) | 0 (0) | 0.36 |
| Distrust in the government | 2 (0.8) | 0 (0) | 1.000 |
| Distrust medical institution | 2 (0.8) | 0 (0) | 1.000 |

<sup>1</sup> Participants who indicated they had not received any COVID-19 vaccine doses were asked to indicate in an open-ended question why they were refusing COVID-19 vaccinations. An individual could indicate $\geq 1$ reason for refusal as such counts may be greater than the total number of individuals.
<sup>2</sup> *p*-value derived from Fisher's exact test

## Discussion

Among participants in a test-negative design case-control study of SARS-CoV-2 infection within California, receipt of seasonal influenza vaccine during the 2021-22 season was strongly associated with receipt of a COVID-19 vaccine and even more so to receiving a booster dose. Older individuals and females were more likely to have received seasonal influenza vaccination, both overall and among the subset of participants who received COVID-19 vaccination. Adults aged 18-49 years were the most likely age group to have received COVID-19 vaccine without seasonal influenza vaccine. While children were more likely than other ages to have received seasonal influenza vaccine only within our study population, this finding was likely driven by the lack of a recommendation for COVID-19 vaccination for children ages 5-11 years until 29 October, 2021 (14). Race and ethnicity were significant predictors of vaccine uptake, with the highest probability of uptake of both vaccines observed among participants identifying as Asian. The highest probability of receiving seasonal influenza vaccine without COVID-19 vaccine was observed among participants who identified as Hispanic, while the highest probability of receiving neither vaccine was observed among participants who identified as Black/African American.

Importantly, participants who tested negative for SARS-CoV-2 in our sample were more likely to have received seasonal influenza vaccine. This finding suggests that unmeasured confounding may pose a risk for future studies aiming to determine the effectiveness of COVID-19 and seasonal influenza vaccines against pathogen-specific endpoints. Behaviors associated with low risk of infection, or with participants’ decision to seek testing at a given threshold of clinical illness, may be closely associated with receipt of multiple vaccines. The finding that an association between negative SARS-CoV-2 test and seasonal influenza vaccination persisted after sub-setting analyses to participants who received COVID-19 vaccination underscores the challenge of controlling for all relevant confounders in future studies (9), particularly those relying only on administrative data capture. Whereas our questionnaire collected behavioral data on risk factors such as mask-wearing and social gatherings, bias in the association between seasonal influenza vaccination and SARS-CoV-2 test outcome persisted even after adjustment for these variables.

Data for our study were collected during the second (2021–2022) of two (2020–2021 and 2021–2022) consecutive seasons with low influenza virus circulation. Uptake of seasonal influenza vaccine nationwide was 44.3% in 2021–2022 (16). Though historically consistent with, or even above coverage levels observed in previous years, 2021–2022 influenza vaccination coverage was lower than population-level estimates of first-dose COVID-19 vaccination coverage and first dose booster coverage (78.1% and 47.3%, respectively) (17, 18). Mitigation strategies for SARS-CoV-2 (use of face masks, social distancing, remote work, avoidance of travel, etc.) likely suppressed transmission of other respiratory viruses including influenza (19). As such, perceived low risk of influenza infection may have influenced lower influenza vaccination uptake as compared to COVID-19 vaccination uptake (20). Avoidance of healthcare facilities where seasonal influenza vaccines are often made available, and other interruptions in routine activities during the COVID-19 pandemic, may have further contributed to differences in seasonal influenza vaccine uptake during the COVID-19 pandemic (21). Given that population-level COVID-19 booster dose coverage mirrors that of seasonal influenza vaccination, and receipt of a COVID-19 booster dose was strongly associated with influenza vaccination, strategies to promote safe co-administration of both COVID-19 and seasonal influenza vaccines may become increasingly important in future respiratory seasons. Clear messaging about the optimal timing of both COVID-19 and influenza vaccination from public health officials and health care providers will likely be important for increasing uptake if both vaccines are recommended seasonally. Efforts to enhance the understanding of reactogenicity of simultaneous administration of both vaccinations must also be prioritized to ensure the safe delivery of both vaccinations, though current estimates suggest co-administration is associated with only mild discomfort.

Our findings that COVID-19 vaccination is associated with influenza vaccine uptake align with studies assessing the impact of the COVID-19 pandemic on influenza vaccine acceptance. A study of vaccine acceptance among undergraduate students in the United States in 2020 found students were more likely to get the COVID-19 vaccine than influenza vaccination, citing social norms as a powerful driver of COVID-19 vaccination (22). However, willingness to receive vaccination is an imperfect predictor of real-world vaccine uptake (23). A key strength of our work is its use of COVID-19 and influenza vaccination uptake, rather than vaccination intentions, and use of a general population sample (24-26).

Our work has limitations. First, we did not adjudicate self-reported vaccination status, which may be influenced by social-desirability biases; however, previous work has identified that misclassification resulting from self-reported vaccination status in telephone surveys is minimal (27). As many workplaces, businesses, and other venues throughout California required individuals to provide proof of COVID-19 vaccination for entry during the study period, we expect that participants were able to report their vaccination status reliably during interviews. Second, our sample is limited to SARS-CoV-2 test seekers, and thus may not be generalizable to populations who may be less likely to seek healthcare; however, wide-spread SARS-CoV-2 testing recommendations and requirements for participation at work, school, or travel throughout the study period likely mitigate this bias. While estimates of COVID-19 and seasonal influenza vaccine uptake within this sample may not reflect population prevalence of vaccine uptake, we are unaware of reasons that associations describing uptake of the two vaccines in relation to each other should not be externally generalizable. Third, given that participants were recruited by telephone, we may have under-sampled certain populations who were unable to answer the phone; of note, the proportion of participants recruited who identified as Hispanic was lower than the proportion of Hispanic individuals residing in the state of California. Fourth, data were not collected on a participants’ intention to receive influenza vaccination. It is possible participants received influenza vaccination or a COVID-19 booster dose after the interview; however, the risk of misclassifying a participants’ seasonal influenza vaccination is low given that 86% of seasonal influenza vaccine doses were administered by 22 October 2021.

COVID-19 vaccine receipt was strongly associated with seasonal influenza vaccine receipt in our study population throughout the 2021–2022 influenza season. Concurrent delivery of both vaccines may be an important strategy to improve coverage of both vaccinations in future respiratory seasons. Benefits of this strategy to maximize vaccine uptake should be weighed against the possible association of vaccine co-administration with increased risk of non-severe adverse events.

## Data Availability

Authors are not authorized to share data.

## Acknowledgements

We would like to thank all participants that gave time to complete our survey making possible this work.

## Disclaimer

The findings and conclusions in this article are those of the author(s) and do not necessarily represent the views or opinions of the California Department of Public Health or the California Health and Human Services Agency.

## Contributions

KLA: Conceived and designed the study and the analysis, wrote the paper

JO: Conceived and designed the study, performed critical review of manuscript

JM: Contributed the data procurement and analysis set curation, performed critical review of manuscript

JW: Conceived and designed the study, performed critical review of manuscript

SJ: Conceived and designed the study, performed critical review of manuscript

JAL: Conceived and designed the study, performed critical review of manuscript

JMP: Conceived and designed the study and the analysis, performed analysis, wrote the paper California COVID-19 Case-Control Study Team: Collected the data

## SUPPLEMENTAL MATERIAL

Dual receipt of COVID-19 and seasonal influenza vaccination uptake in California (USA) during the 2021-2022 influenza season

## Table of Contents

**Table S1.**
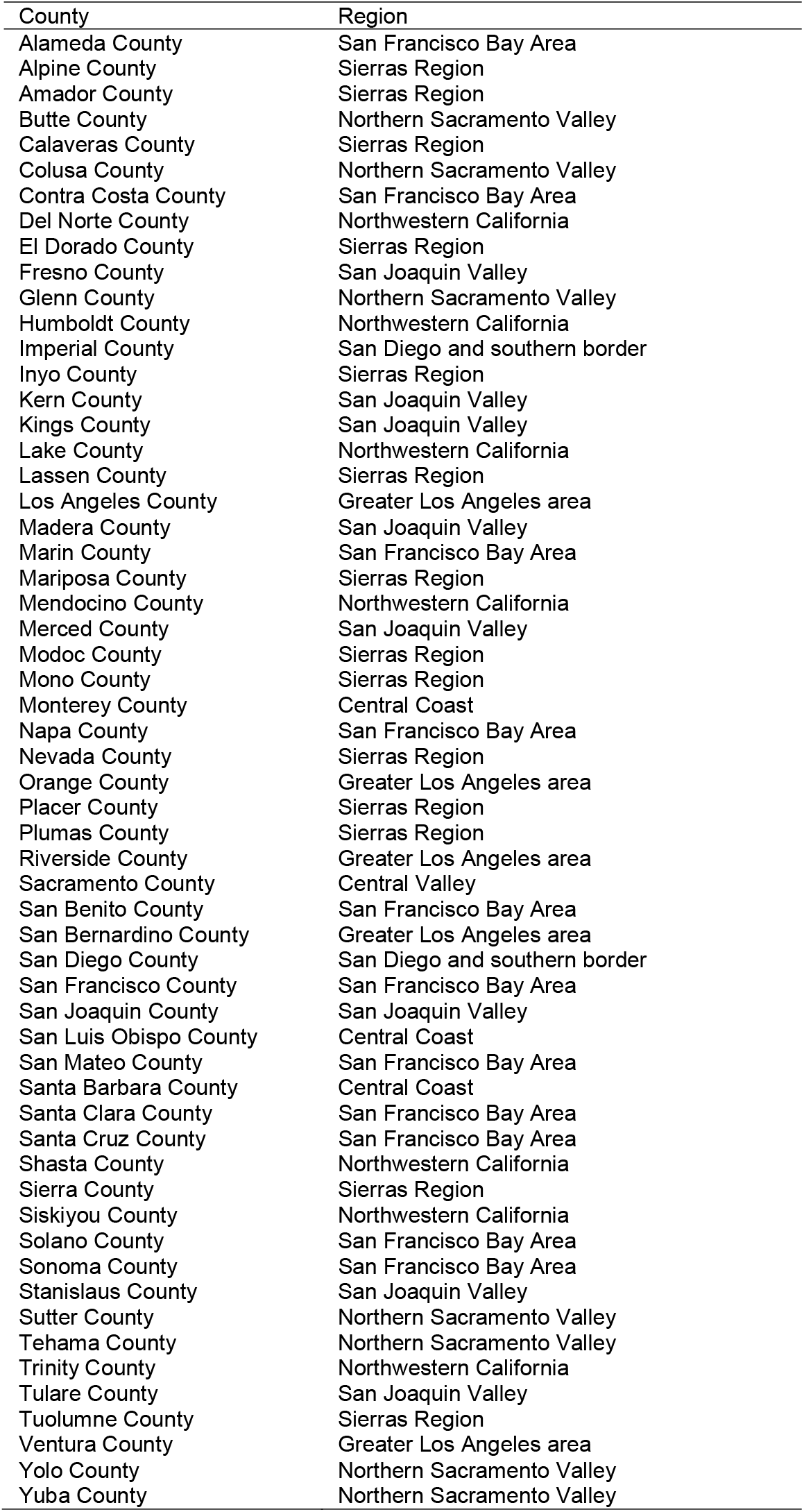
Counties included in each geographic region.

| County | Region |
| --- | --- |
| Alameda County | San Francisco Bay Area |
| Alpine County | Sierras Region |
| Amador County | Sierras Region |
| Butte County | Northern Sacramento Valley |
| Calaveras County | Sierras Region |
| Colusa County | Northern Sacramento Valley |
| Contra Costa County | San Francisco Bay Area |
| Del Norte County | Northwestern California |
| El Dorado County | Sierras Region |
| Fresno County | San Joaquin Valley |
| Glenn County | Northern Sacramento Valley |
| Humboldt County | Northwestern California |
| Imperial County | San Diego and southern border |
| Inyo County | Sierras Region |
| Kern County | San Joaquin Valley |
| Kings County | San Joaquin Valley |
| Lake County | Northwestern California |
| Lassen County | Sierras Region |
| Los Angeles County | Greater Los Angeles area |
| Madera County | San Joaquin Valley |
| Marin County | San Francisco Bay Area |
| Mariposa County | Sierras Region |
| Mendocino County | Northwestern California |
| Merced County | San Joaquin Valley |
| Modoc County | Sierras Region |
| Mono County | Sierras Region |
| Monterey County | Central Coast |
| Napa County | San Francisco Bay Area |
| Nevada County | Sierras Region |
| Orange County | Greater Los Angeles area |
| Placer County | Sierras Region |
| Plumas County | Sierras Region |
| Riverside County | Greater Los Angeles area |
| Sacramento County | Central Valley |
| San Benito County | San Francisco Bay Area |
| San Bernardino County | Greater Los Angeles area |
| San Diego County | San Diego and southern border |
| San Francisco County | San Francisco Bay Area |
| San Joaquin County | San Joaquin Valley |
| San Luis Obispo County | Central Coast |
| San Mateo County | San Francisco Bay Area |
| Santa Barbara County | Central Coast |
| Santa Clara County | San Francisco Bay Area |
| Santa Cruz County | San Francisco Bay Area |
| Shasta County | Northwestern California |
| Sierra County | Sierras Region |
| Siskiyou County | Northwestern California |
| Solano County | San Francisco Bay Area |
| Sonoma County | San Francisco Bay Area |
| Stanislaus County | San Joaquin Valley |
| Sutter County | Northern Sacramento Valley |
| Tehama County | Northern Sacramento Valley |
| Trinity County | Northwestern California |
| Tulare County | San Joaquin Valley |
| Tuolumne County | Sierras Region |
| Ventura County | Greater Los Angeles area |
| Yolo County | Northern Sacramento Valley |
| Yuba County | Northern Sacramento Valley |

**Table S2.**
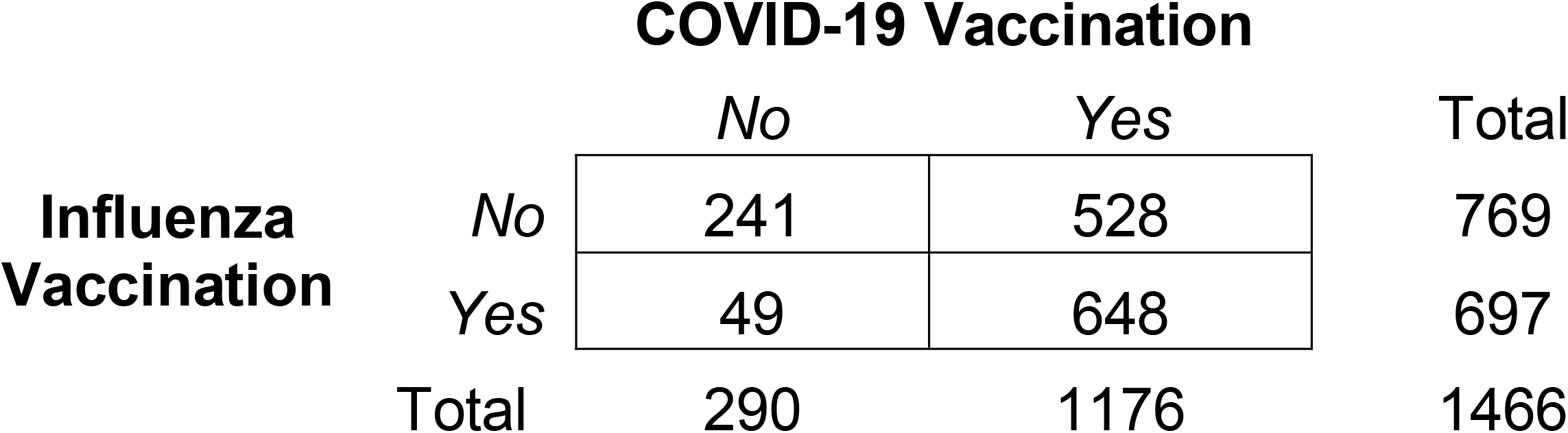
2×2 table for influenza and COVID-19 vaccination

**Table S3.** Characteristics of participants by COVID-19 vaccination status.

|  |  | No COVID-19 vaccines<br>(%)<br>N = 290 | Primary series<br>N (%)<br>N = 460 | ≥1 booster dose<br>N (%)<br>N = 511 |
| --- | --- | --- | --- | --- |
| Birth Sex | Female | 138 (47.6) | 248 (54.0) | 331 (64.8) |
|  | Male | 152 (52.4) | 211 (46.0) | 180 (35.2) |
| Age Category | 5-11 years | 69 (23.8) | 40 (8.7) | 0 (0.0) |
|  | 12-17 years | 36 (12.4) | 24 (5.2) | 11 (2.2) |
|  | 18-29 years | 48 (16.6) | 123 (26.7) | 140 (27.4) |
|  | 30-49 years | 79 (27.2) | 168 (36.5) | 175 (34.2) |
|  | 50-64 years | 38 (13.1) | 73 (15.9) | 93 (18.2) |
|  | 65+ years | 20 (6.9) | 32 (7.0) | 92 (18.0) |
| Race/Ethnicity | White | 114 (39.3) | 233 (50.7) | 273 (53.4) |
|  | Asian | 17 (5.9) | 72 (15.7) | 94 (18.4) |
|  | Black/African American | 21 (7.2) | 16 (3.5) | 13 (2.5) |
|  | Hispanic | 77 (26.6) | 90 (19.6) | 90 (17.6) |
|  | Native American/Native Hawaiian | 7 (2.4) | 7 (1.5) | 6 (1.2) |
|  | Multiracial | 40 (13.8) | 34 (7.4) | 29 (5.7) |
|  | Refused to disclose | 14 (4.8) | 8 (1.7) | 6 (1.2) |
| Reported Comorbidity | No co-morbidity | 226 (78.2) | 338 (73.6) | 335 (65.8) |
|  | ≥1 co-morbidity | 63 (21.8) | 121 (26.4) | 174 (34.2) |
| Region | San Francisco Bay Area | 12 (4.1) | 54 (11.7) | 80 (15.7) |
|  | Central Coast | 24 (8.3) | 51 (11.1) | 72 (14.1) |
|  | Greater Sacramento Region | 34 (11.7) | 55 (12.0) | 68 (13.3) |
|  | Northern Sacramento Valley | 32 (11.0) | 43 (9.3) | 67 (13.1) |
|  | San Joaquin Valley | 52 (17.9) | 47 (10.2) | 43 (8.4) |
|  | Northwestern California | 32 (11.0) | 64 (13.9) | 44 (8.6) |
|  | Sierras Region | 28 (9.7) | 41 (8.9) | 41 (8.0) |
|  | San Diego and Southern Border | 31 (10.7) | 55 (12.0) | 53 (10.4) |
|  | Greater Los Angeles Area | 45 (15.5) | 50 (10.9) | 43 (8.4) |
| SARS-CoV-2 Infection Status | SARS-CoV-2 negative (control) | 131 (45.2) | 200 (43.5) | 292 (57.1) |
|  | SARS-CoV-2 positive (case) | 159 (54.8) | 260 (56.5) | 219 (42.9) |
| Use of face masks | No mask use in public settings | 60 (20.7) | 46 (10.0) | 54 (10.6) |
|  | Mask use in public settings | 230 (79.3) | 414 (90.0) | 456 (89.4) |
| Attended public setting | Did not attend gathering | 155 (53.8) | 200 (43.5) | 198 (38.9) |
|  | Attended gathering | 133 (46.2) | 260 (56.5) | 311 (61.1) |
| Month of Interview | October (2021) | 11 (3.8) | 27 (5.9) | 5 (1.0) |
|  | November (2021) | 49 (16.9) | 90 (19.6) | 19 (3.7) |
|  | December (2021) | 83 (28.6) | 104 (22.6) | 55 (10.8) |
|  | January (2022) | 65 (22.4) | 102 (22.2) | 106 (20.7) |
|  | February (2022) | 21 (7.2) | 34 (7.4) | 59 (11.5) |
|  | March (2022) | 22 (7.6) | 27 (5.9) | 61 (11.9) |
|  | April (2022) | 17 (5.9) | 27 (5.9) | 95 (18.6) |
|  | May (2022) | 12 (4.1) | 32 (7.0) | 68 (13.3) |
|  | June (2022) | 10 (3.4) | 17 (3.7) | 43 (8.4) |

**Table S4.** Characteristics of participants receiving one or two booster doses following the primary series COVID-19 vaccination.

|  |  | **1 booster dose** | **2 booster doses** |
| --- | --- | --- | --- |
|  |  | N (%)<br>N = 490 | N (%)<br>N = 21 |
| Birth Sex | Female | 316 (64.5) | 15 (71.4) |
|  | Male | 174 (35.5) | 6 (28.6) |
| Age Category | 5-11 years | 0 (0.0) | 0 (0.0) |
|  | 12-17 years | 11 (2.2) | 0 (0.0) |
|  | 18-29 years | 139 (28.4) | 1 (4.8) |
|  | 30-49 years | 174 (35.5) | 1 (4.8) |
|  | 50-64 years | 85 (17.3) | 8 (38.1) |
|  | 65+ years | 81 (16.5) | 11 (52.4) |
| Race/Ethnicity | White | 29 (5.9) | 0 (0.0) |
|  | Asian | 93 (19.0) | 1 (4.8) |
|  | Black/African American | 13 (2.7) | 0 (0.0) |
|  | Hispanic | 89 (18.2) | 1 (4.8) |
|  | Native American/Native Hawaiian | 6 (1.2) | 0 (0.0) |
|  | Multiracial | 6 (1.2) | 0 (0.0) |
|  | Refused to disclose | 254 (51.8) | 19 (90.5) |
| Reported Comorbidity | No co-morbidity | 327 (66.9) | 8 (40.0) |
|  | ≥1 co-morbidity | 162 (33.1) | 12 (60.0) |
| Region | San Francisco Bay Area | 77 (15.7) | 3 (14.3) |
|  | Central Coast | 69 (14.1) | 3 (14.3) |
|  | Greater Sacramento Region | 64 (13.1) | 4 (19.0) |
|  | Northern Sacramento Valley | 64 (13.1) | 3 (14.3) |
|  | San Joaquin Valley | 42 (8.6) | 1 (4.8) |
|  | Northwestern California | 43 (8.8) | 1 (4.8) |
|  | Sierras Region | 36 (7.3) | 5 (23.8) |
|  | San Diego and Southern Border | 53 (10.8) | 0 (0.0) |
|  | Greater Los Angeles Area | 42 (8.6) | 1 (4.8) |
| SARS-CoV-2 Infection Status | SARS-CoV-2 negative (control) | 51 (10.4) | 3 (14.3) |
|  | SARS-CoV-2 positive (case) | 438 (89.6) | 18 (85.7) |
| Use of face masks | No mask use in public settings | 191 (39.1) | 7 (33.3) |
|  | Mask use in public settings | 297 (60.9) | 14 (66.7) |
| Attended public setting | Did not attend gathering | 282 (57.6) | 10 (47.6) |
|  | Attended gathering | 208 (42.4) | 11 (52.4) |
| Month of Interview | October (2021) | 5 (1.0) | 0 (0.0) |
|  | November (2021) | 19 (3.9) | 0 (0.0) |
|  | December (2021) | 55 (11.2) | 0 (0.0) |
|  | January (2022) | 106 (21.6) | 0 (0.0) |
|  | February (2022) | 59 (12.0) | 0 (0.0) |
|  | March (2022) | 61 (12.4) | 0 (0.0) |
|  | April (2022) | 91 (18.6) | 4 (19.0) |
|  | May (2022) | 59 (12.0) | 9 (42.9) |
|  | June (2022) | 35 (7.1) | 8 (38.1) |

**Table S5.** Reasons for SARS-CoV-2 testing stratified by influenza and COVID-19 vaccine receipt and SARS-CoV-2 infection status.

| *SARS-CoV-2 infection* | **Neither vaccine** |  | **Influenza vaccine only** |  | **COVID-19 vaccine only<sup>2</sup>** |  | **Both vaccines** |  |
| --- | --- | --- | --- | --- | --- | --- | --- | --- |
|  | *Positive*<br>*n (%)*<br>*N=135* | *Negative*<br>*n (%)*<br>*N=106* | *Positive*<br>*n (%)*<br>*N=24* | *Negative*<br>*n (%)*<br>*N=25* | *Positive*<br>*n (%)*<br>*N=245* | *Negative*<br>*n (%)*<br>*N=181* | *Positive*<br>*n (%)*<br>*N=234* | *Negative*<br>*n (%)*<br>*N=311* |
| Reason for SARS-CoV-2 Testing<sup>1</sup> |  |  |  |  |  |  |  |  |
| Curious to see if infected | 24 (17.8) | 13 (12.3) | 4 (16.7) | 30 (16.6) | 37 (15.1) | 30 (16.6) | 48 (20.5) | 58 (18.6) |
| Travel requirement | 1 (0.7) | 8 (7.5) | 0 (0.0) | 14 (7.7) | 9 (3.7) | 14 (7.7) | 8 (3.4) | 19 (6.1) |
| Experiencing COVID-19 symptoms | 79 (58.5) | 20 (18.9) | 15 (62.5) | 38 (21.0) | 160 (65.3) | 38 (21.0) | 140 (59.8) | 48 (15.4) |
| Screening required | 16 (11.9) | 41 (38.7) | 0 (0.0) | 50 (27.6) | 16 (6.5) | 50 (27.6) | 23 (9.8) | 105 (33.8) |
| Medical procedure | 5 (3.7) | 16 (15.1) | 3 (12.5) | 27 (14.9) | 4 (1.6) | 27 (14.9) | 10 (4.3) | 56 (18.0) |
| Public health recommendation to get tested | 1 (0.7) | 1 (0.9) | 0 (0.0) | 1 (0.6) | 3 (1.2) | 1 (0.6) | 4 (1.7) | 0 (0.0) |
| Household COVID-19 contact | 7 (5.2) | 5 (4.7) | 1 (4.2) | 5 (2.8) | 7 (2.9) | 5 (2.8) | 11 (4.7) | 4 (1.3) |
| Other contact with COVID-19 | 45 (33.3) | 12 (11.3) | 10 (41.7) | 40 (22.1) | 73 (29.8) | 40 (22.1) | 72 (30.8) | 62 (19.9) |

<sup>1</sup>Since interviewers indicated all reasons listed by participants, and some participants refused to respond to the question, reasons will not sum to the total sample size.
<sup>2</sup>Participants were categorized as vaccinated for COVID-19 if they had received $\geq 1$ dose of BNT162b2 [Pfizer/BioNTech], mRNA-1273 [Moderna], or Ad26.COV2.S [Janssen] at the time of their interview

**Figure S1.**
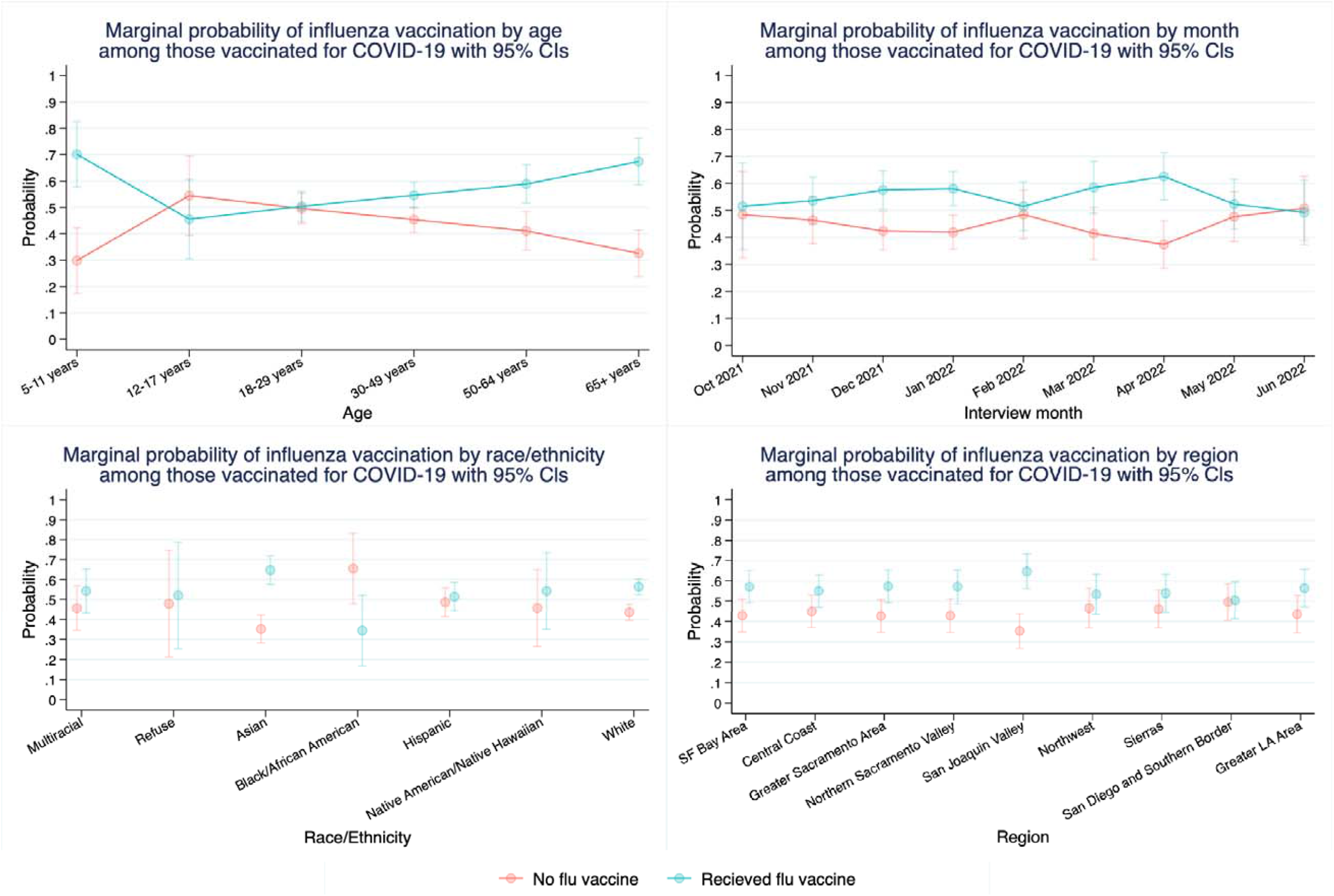
Margins plots among participants vaccinated for COVID-19. A) marginal probability of vaccination status by age B) marginal probability of vaccination status by SARS-CoV-2 test month associated with study enrollment C) marginal probability of vaccination status by race/ethnicity D) marginal probability of vaccination status by region Note: CI – confidence interval.

